# Dynamic frame-by-frame motion correction for ^18^F-flurpiridaz PET-MPI using convolution neural network

**DOI:** 10.1101/2025.06.27.25330436

**Authors:** Meghana Urs, Aditya Killekar, Valerie Builoff, Mark Lemley, Chih-Chun Wei, Giselle Ramirez, Paul Kavanagh, Christopher Buckley, Piotr J. Slomka

## Abstract

**Purpose:** Precise quantification of myocardial blood flow (MBF) and flow reserve (MFR) in 18F-flurpiridaz PET significantly relies on motion correction (MC). However, the manual frame-by-frame correction leads to significant inter-observer variability, time-consuming, and requires significant experience. We propose a deep learning (DL) framework for automatic MC of 18F-flurpiridaz PET.

**Methods:** The method employs a 3D ResNet based architecture that takes 3D PET volumes and outputs motion vectors. It was validated using 5-fold cross-validation on data from 32 sites of a Phase III clinical trial (NCT01347710). Manual corrections from two experienced operators served as ground truth, and data augmentation using simulated vectors enhanced training robustness. The study compared the DL approach to both manual and standard non-AI automatic MC methods, assessing agreement and diagnostic accuracy using minimal segmental MBF and MFR.

**Results:** The area under the receiver operating characteristic curves (AUC) for significant CAD were comparable between DL-MC MBF, manual-MC MBF from Operators (AUC=0.897,0.892 and 0.889, respectively; p>0.05), standard non-AI automatic MC (AUC=0.877; p>0.05) and significantly higher than No-MC (AUC=0.835; p<0.05). Similar findings were observed with MFR. The 95% confidence limits for agreement with the operator were ±0.49ml/g/min (mean difference = 0.00) for MFR and ±0.24ml/g/min (mean difference = 0.00) for MBF.

**Conclusion:** DL-MC is significantly faster but diagnostically comparable to manual-MC. The quantitative results obtained with DL-MC for MBF and MFR are in excellent agreement with those manually corrected by experienced operators compared to standard non-AI automatic MC in patients undergoing ^18^F-flurpiridaz PET-MPI.

## Introduction

Positron emission tomography (PET) myocardial perfusion imaging (MPI) has demonstrated improved diagnostic and prognostic accuracy for coronary artery disease (CAD) compared to other imaging modalities (*1,2*). Studies have shown that quantification of myocardial blood flow (MBF) and myocardial flow reserve (MFR) using dynamic PET offers enhanced diagnostic value over conventional relative MPI (*3*). Motion correction (MC) is particularly crucial for accurately quantifying MBF and MFR from dynamic PET-MPI data (*4*), as patient motion – including cardiac, respiratory, and body movement during scanning – can introduce error in MBF estimation (*5,6*).

In dynamic cardiac PET imaging, a sequence of images is continuously taken over several minutes after injecting a radio-labeled tracer. This allows visualization of how the tracer spreads through and is absorbed by the heart muscle as blood perfusion occurs. ^18^F-flurpiridaz is a novel PET-MPI tracer that is anticipated to see wide-spread clinical adoption. Our recent research has highlighted the importance of residual activity correction (RAC) and manual as well as automatic frame-by-frame MC in analyzing dynamic PET flow in same-day rest/stress studies using ^18^F-flurpiridaz (*7,8*).

One of our prior works involved manual frame-by-frame corrections (*7*), however, this method is time-intensive and leads to inter-observer variability. Data-driven motion detection methods (*9,10*), which do not require external devices, are favored for their potential to streamline clinical motion correction implementation. Nevertheless, there is a scarcity of research focused on correcting inter-frame motion in ^18^F-flurpiridaz cardiac dynamic PET imaging. Previous work reported an automated algorithm based on aligning each frame to a static model of the ventricles defined by automatically segmented left and right ventricle contours (*8*). Although such an approach is significantly faster compared to manual-MC, it produced some outliers as compared to experienced operators. There is a further need to enhance the performance of such pre-processing tools to optimize overall cardiac PET processing.

Deep learning (DL) has exhibited promising capabilities across a wide range of medical imaging tasks, including image segmentation, image registration, image generation and enhancement. The use of deep learning for motion detection in sequential images is still in its early stages. Shi et al. introduced a deep learning-based approach for the automated correction of motion artifacts in dynamic cardiac Rubidium-82 chloride PET imaging (*11*). However, only generic simulated motion was utilized for evaluating the network, and the results were not clinically evaluated for agreement with experienced operators or diagnostic accuracy. To optimize the clinical utility of ^18^F-flurpiridaz, we propose a novel deep learning framework for automatic MC of ^18^F-flurpiridaz PET-MPI using 3D Resnet based architecture using data from the initial phase III Flurpiridaz trial (NCT01347710) and manual-MC vectors from experienced operators as ground truth. To enhance training robustness and expand the dataset, we implemented data augmentation by generating simulated motion vectors from bootstrap distributions of these manual corrections.

## Materials and Methods

### Study Population

The Phase III Flurpiridaz PET trial (flurpiridaz-301) involved 795 patients diagnosed with or suspected of having CAD. The trial was conducted across 72 clinical sites in the United States, Canada, and Finland from 2011 to 2013. Among these patients, 755 had evaluable rest-stress Flurpiridaz PET-MPI, rest-stress SPECT-MPI, and quantitative invasive coronary angiography (*12*). Dynamic imaging data was available for 275 of the 755 evaluable patients. However, 44 of these patients were excluded due to missing or corrupted dynamic images, issues with the left ventricle input curve, or unavailable heart rate and blood pressure data at rest. Thus, 231 patients from 32 different sites had suitable data for network training and quantitative blood flow analysis. Details on the trial participants is presented in Supplemental Methods (*12*).

### PET protocol

PET imaging was performed using PET scanners in 2D or 3D mode after participants fasted for a minimum of 3 hours. A detailed description can be found in Supplemental Methods.

### Manual motion correction

In both stress and rest phases, motion correction for each frame was performed in 1 mm increments by two experienced operators. Their task was to align myocardial tracer uptake precisely with myocardial contours, which are derived from the static part of the dynamic sequence (*7,13*). Specifically, for every frame within each dataset, the operators adjusted the image to match the static left ventricle myocardial contours along three principal axes: septal-lateral, anterior-inferior, and apical-basal directions (*7*).

### Standard clinical automatic motion correction

The automatic MC algorithm (*8*) is based on aligning each frame to a static model of the ventricles defined by automatically segmented left and right ventricle contours. A more detailed explanation is presented in Supplemental Methods.

### Deep learning-based automatic motion correction

The goal of the algorithm was to align all the frames of a dynamic PET scan to a reference frame. To achieve this, a neural network was trained to predict the translation vector (t_x_, t_y_, t_z_), accounting for the motion between a given and reference frame. To predict the motion vector required to align the given frame with the reference frame, the neural network required a dual-channel input to correlate features between them. Our network shown in Figure 1 adapts a 3D ResNet (*14*) based architecture with a regression head at the end to predict translation vectors (t_x_, t_y_, t_z_). The ResNet 3D convolution (Conv3D) blocks extract the feature vectors from both given and reference frames separately in the initial layers which are then concatenated and parsed through future Conv3D layers. A comprehensive description of the architecture and loss functions can be found in Supplemental Methods.

**FIG. 1.**
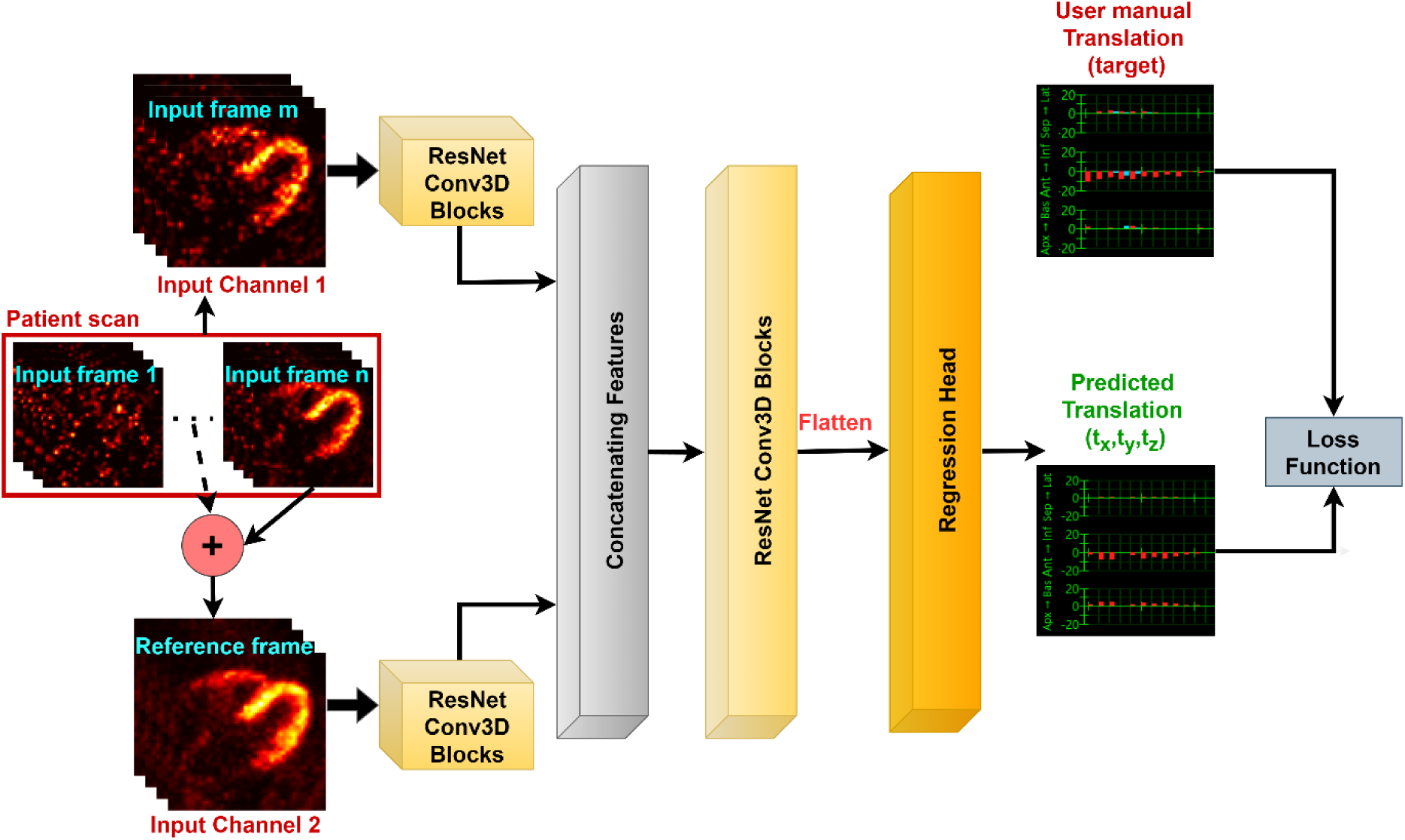
Proposed architecture for motion translation vector prediction. Input channel 1 represents the given frame (m) and channel 2 is the reference frame, intensity summed frames with 120s<t<3200s. Conv3D: 3D Convolution, t_x_: translation x direction, t_y_: translation y direction, t_z_: translation z direction, f(m): function of frame number m

### Training dataset for AI motion correction

Intensity summed frames with acquisition time 120s<t<3200s were used as reference frames. Target/ground truth was an operator manual corrected motion translation vector. Since the motion correction vectors were from two different operators, a randomizer was used to pick one target vector per case.

To enhance the robustness of the training process and significantly expand the dataset, an advanced strategy of data augmentation was employed. This involved generating a multitude of simulated motion vectors derived from the bootstrap distributions of the original manual corrections per frame as shown in Figure 2. Bootstrapping technique treats the original sample of manual operator corrections as a small representation of a much larger, unknown *true* distribution of possible movements. New samples are created by repeatedly drawing motion vector values with replacement from that original sample. By repeating this thousands of times underlying distribution of likely movements for that frame can be estimated. It captures the central tendency and variability of the original expert operator corrections. By creating these synthetic yet realistic motion scenarios, that are statistically similar to the original expert corrections, the model was exposed to a much broader spectrum of potential patient movements, which in turn improved its ability to accurately correct for motion in diverse clinical settings and reduced the risk of overfitting to the limited initial manual dataset. A detailed explanation on the dataset and data pre-processing can be found in Supplemental Methods.

**FIG. 2.**
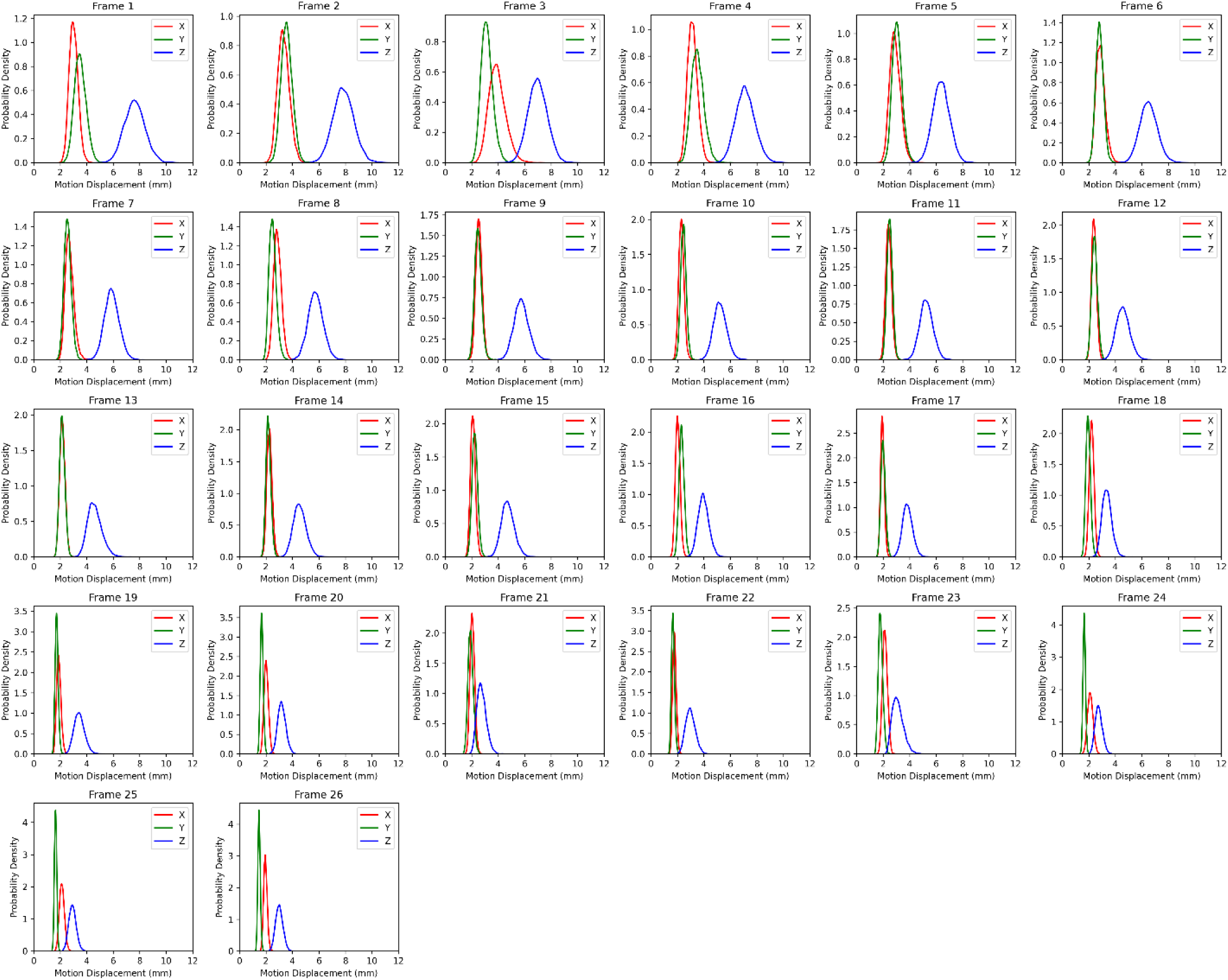
Bootstrap probability density distributions of the original manual motion displacement corrections per frame (in mm) of the dynamic PET scans in X (red), Y (green) and Z (blue) directions. Simulated motion vectors that emulate real patient motion are derived from these distributions for data augmentation

### Training and Testing regimen

Five-fold cross-validation was used for a rigorous, unbiased evaluation of the framework. This method utilized five separate models, each trained independently, along with exclusive hold-out (test) sets, each comprising 20% of the total dataset of 231 cases from 32 different sites. To make testing more thorough, inference was run on an external dataset, ensuring no sites overlapped between the training and test sets to improve generalization. The entire cohort was divided into five mutually exclusive subsets for this purpose, with each subset containing cases (46-47 patients) from distinct sites (Figure 3). Details about 5-fold cross-validation and training parameters used can be found in Supplemental Methods (*15*).

**FIG. 3.**
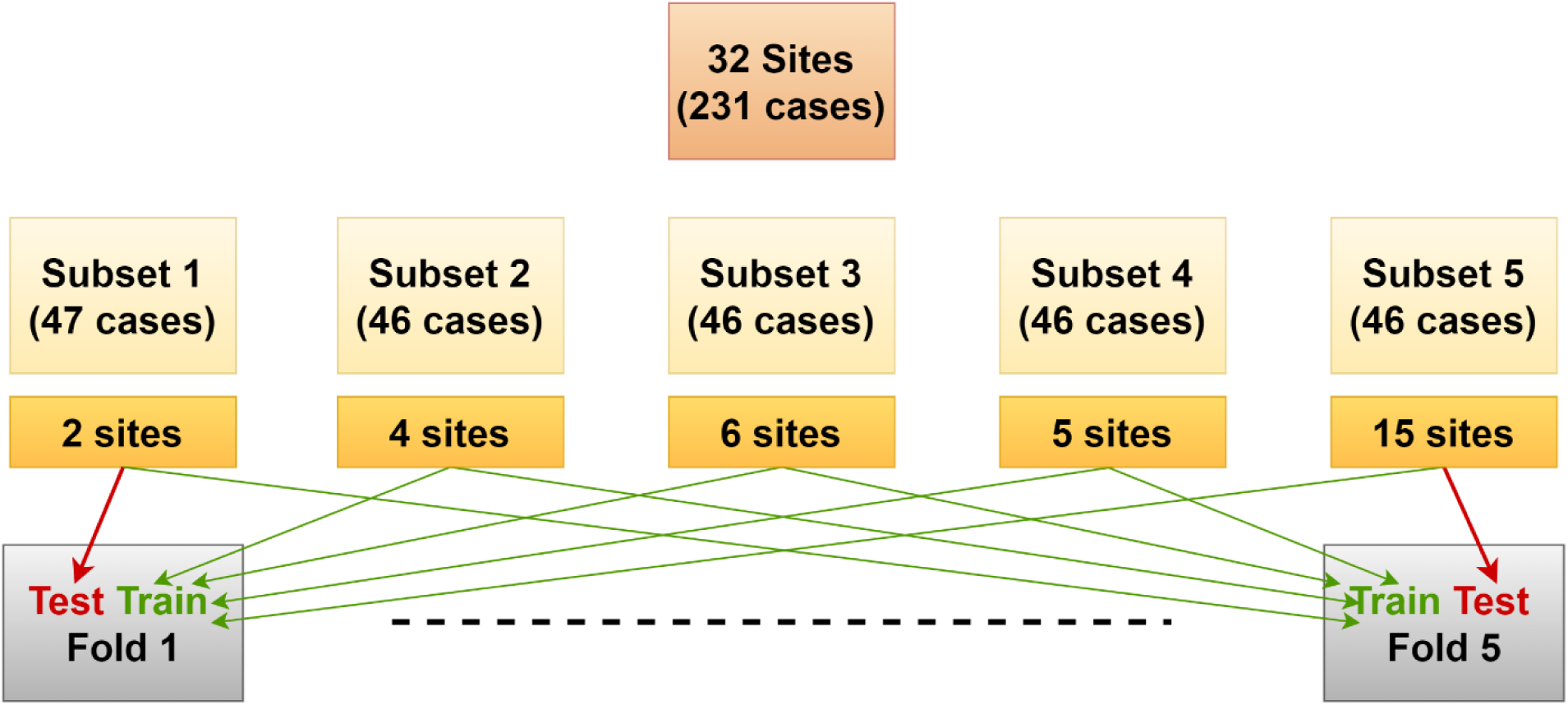
Site wise splitting of dataset to achieve five-fold external validation

### Myocardial blood flow and myocardial flow reserve quantification

To process dynamic PET, and compute MBF and MFR, we used QPET software (Cedars-Sinai, Los Angeles, California, United States) by applying the 2-compartment kinetic model proposed by Packard et al. (*16*). A detailed description of MBF and MFR quantification methods is provided in the Supplemental Methods (*4,7,17–19*).

### Quantitative Evaluation

The motion vector prediction was evaluated in terms of translation root mean square error (RMSE) of all the frames in the test data. The average error was calculated as 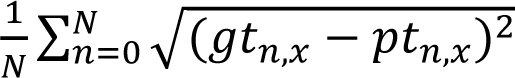, 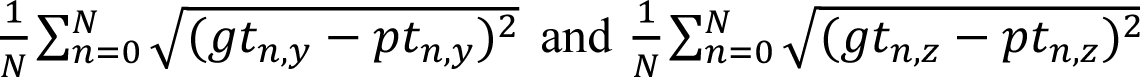, for x, y and z directions respectively, where (*gt_n,x_*, *gt_n,y_*, *gt_n,z_*) and (*pt_n,x_*, *pt_n,y_*, *pt_n,z_*) are the motion ground-truth and prediction motion translation vectors in three directions for frame *n*, and *N* = is the total number of frames in the test dataset. The final error was the average error over the external test sets across all 5 folds.

### Statistical Analysis

Performance of the DL-MC is demonstrated using Bland-Altman (BA) and correlation plots. Concordance correlation coefficient (CCC) between DL-MC and avg manual-MC MBF and MFR were calculated to assess the agreement between the two methods. Lin’s CCC was calculated by measuring the variation of the linear relationship between each pair of data from a line through the origin (*20*). CCCs are shown between DL-MC and avg manual-MC MBF and MFR per segment. Comparison of the agreement with experienced operators of the conventional automated method and DL-MC was performed with Pitman-Morgan test of paired variance (*21,22*) using R studio version 2024.04.2. Lin’s CCC was compared by calculating the z-scores using Fisher’s z-transformation (*23*) and assessing for statistically significant differences between the z-values.

Comparison between manual-MC, DL-MC, standard non-AI automatic MC and no-MC was conducted by analyzing the diagnostic performance of stress MBF and MFR using areas under the receiver operating characteristic curve (AUC) as per DeLong et al. (*24*). A p-value less than 0.05 was considered statistically significant.

## Results

### Patient Population

Patient characteristics are presented in Table 1. Out of 231 patients, 51 had obstructive CAD, of whom 44 were male. The mean age was 62 years with a standard deviation of 9 years.

**TABLE 1.** Patient Characteristics

|  | **Overall** | **Obstructive CAD** | **No Obstructive CAD** | **P-Value** |
| --- | --- | --- | --- | --- |
| **Number of Patients, n (%)** | 231 | 51 (22%) | 180 (78%) |  |
| **Age (years), mean (SD)** | 62 (9) | 64 (10) | 61 (9) | 0.058 |
| **Biological Sex, n (%)** |  |  |  | **0.006** |
| **Male** | 161 (70%) | 44 (86%) | 117 (65%) |  |
| **Female** | 70 (30%) | 7 (14%) | 63 (35%) |  |
| **Race, n (%)** |  |  |  | 0.994 |
| **White** | 179 (77%) | 39 (76%) | 140 (78%) |  |
| **Other** | 52 (23%) | 12 (24%) | 40 (22%) |  |
| **Ethnicity, n (%)** |  |  |  | 0.402 |
| **Hispanic** | 10 (4%) | 3 (6%) | 7 (4%) |  |
| **Not Hispanic** | 210 (91%) | 44 (86%) | 166 (92%) |  |
| **Not Reported** | 11 (5%) | 4 (8%) | 7 (4%) |  |
| **BMI (kg/m<sup>2</sup>), median [Q1, Q3]** | 31 [27,35] | 30 [27,34] | 31 [27,36] | 0.339 |
| **Family History of CAD, n (%)** | 129 (56%) | 27 (53%) | 102 (57%) | 0.295 |
| **Hypertension, n (%)** | 199 (86%) | 43 (84%) | 156 (87%) | 0.842 |
| **Hyperlipidemia, n (%)** | 202 (87%) | 45 (88%) | 157 (87%) | 1.000 |
| **History of prior MI, n (%)** | 36 (16%) | 11 (22%) | 25 (14%) | 0.246 |
| **Diabetes, n (%)** | 83 (36%) | 17 (33%) | 66 (37%) | 0.785 |
| **Smoking, n (%)** | 135 (58%) | 32 (63%) | 103 (57%) | 0.694 |
| **History of prior PCI, n (%)** | 74 (32%) | 15 (29%) | 59 (33%) | 0.773 |

CAD = coronary artery disease, BMI = body mass index, MI = myocardial infarction, PCI = percutaneous coronary intervention, SD = standard deviation. Significant p-values are shown in bold.

### Speed of Motion Correction

Average time taken for manual-MC was about 10 minutes per patient. Standard non-AI automatic MC takes <10 seconds to perform motion correction (*8*). Inference time for our trained neural network is about 1 second with GPU and 1.5 seconds with CPU per patient scan.

### Quantitative evaluation of motion correction methods

Average RMSE and the standard deviation across all 5 folds were significantly lower for DL-MC in comparison to non-AI automatic MC. The average maximum error in each direction also showed considerable improvement. A detailed summary of the evaluation results is presented in Table 2.

**TABLE 2.**
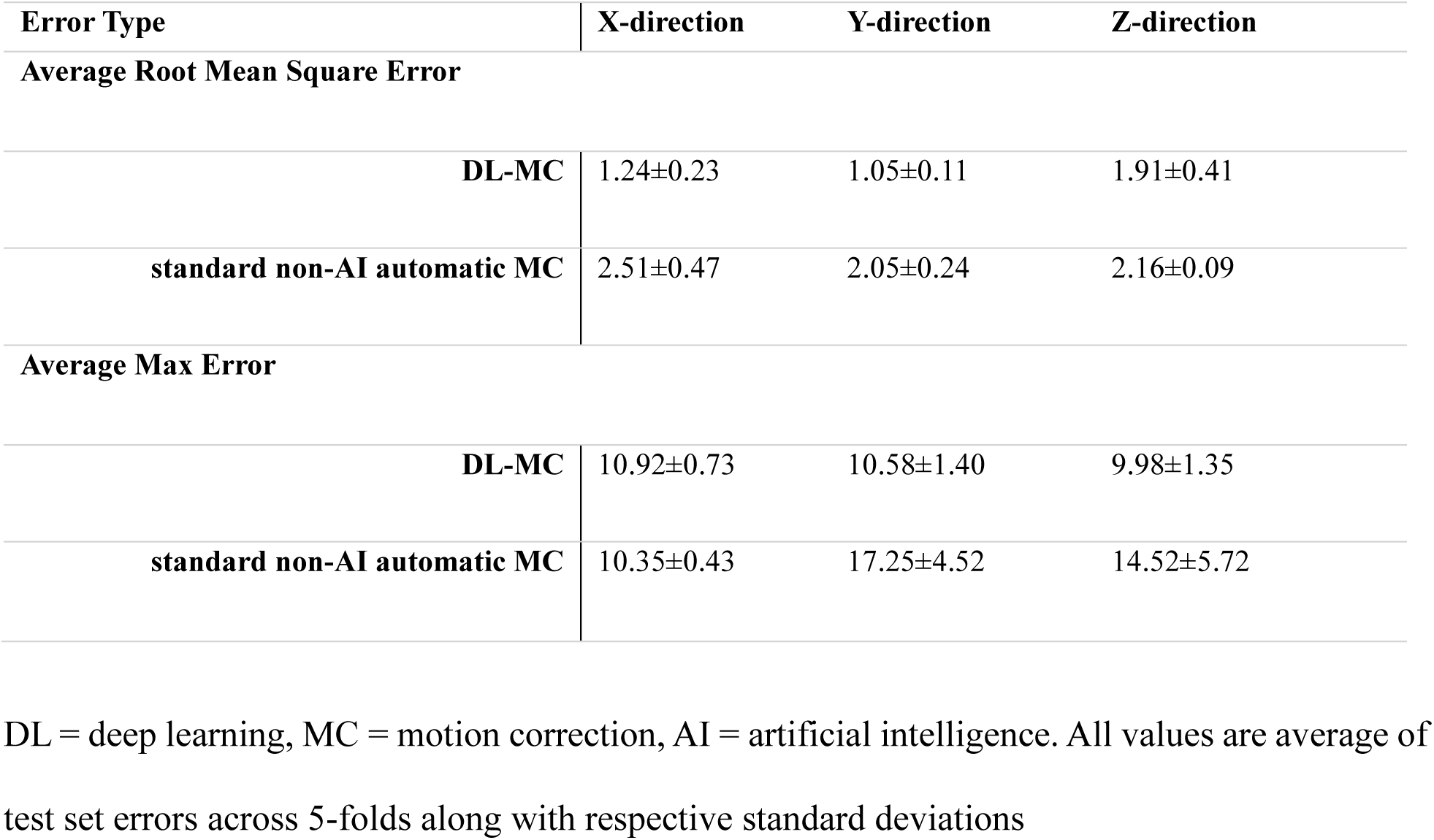
Quantitative evaluation results

### Agreement with Operators

#### Myocardial Flow Reserve

Figure 4a shows BA and correlation plots for standard non-AI automatic MC and operator average MC. Figure 4b shows the BA and correlation plots between the DL-MC and the average MC by two experienced operators for MFR. The variability around the mean difference for DL-MC, ±0.49 (mean difference = 0.00), was lower than variability for non-AI automatic MC, ±0.82 (mean difference = -0.02). Morgan-Pitman resulted in statistically significant difference in variances, with a p-value <0.00001. The bottom row shows correlations between minimal segmental MFR with average of manual-MC from two operators and deep learning based automatic MC with CCC=0.943 (95% CI: 0.900, 0.986) and non-AI automatic MC with CCC=0.864 (95% CI: 0.798, 0.929). Fisher’s z-score evaluation resulted in statistically significant difference in CCC, p-value <0.00001.

**FIG. 4.**
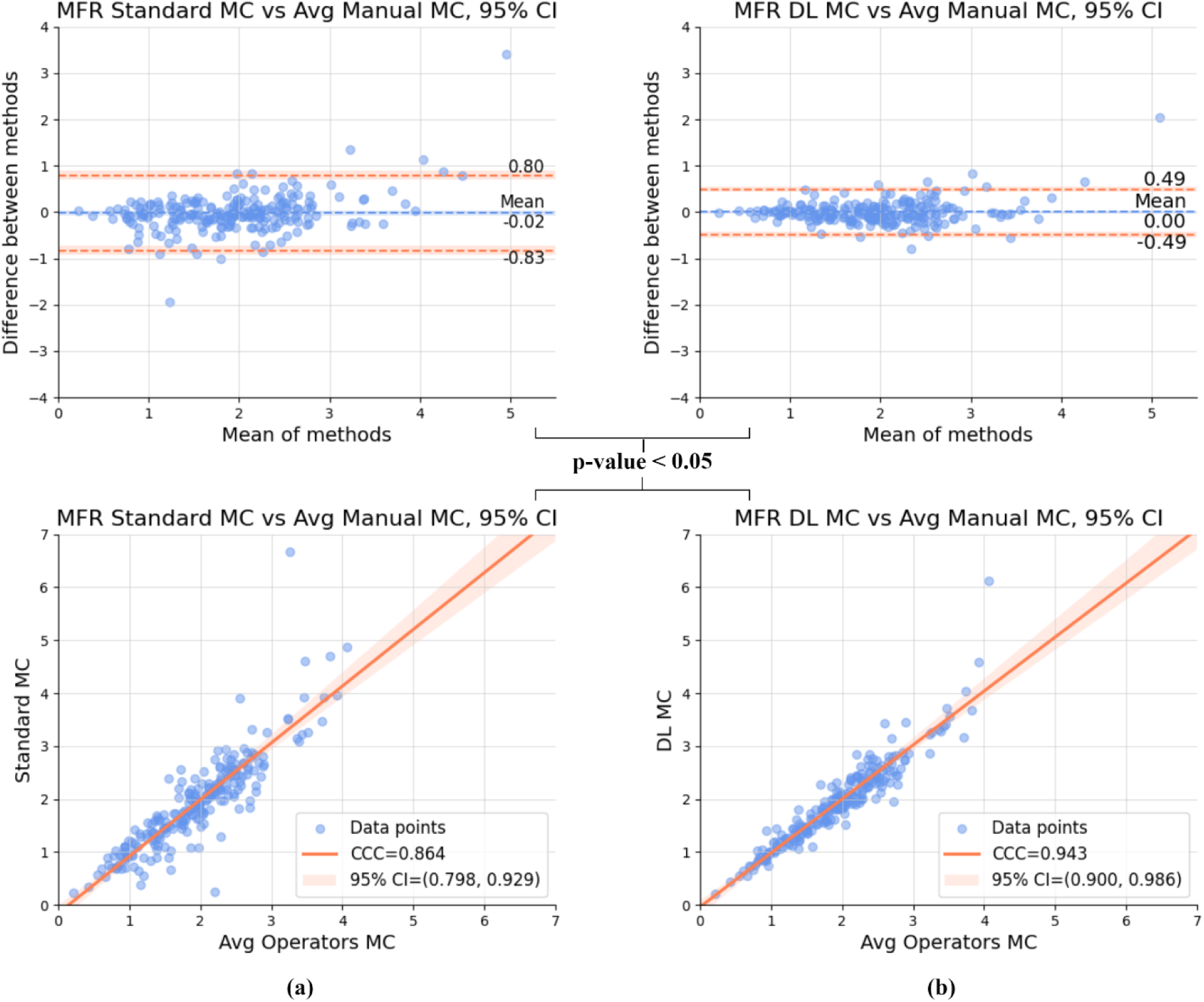
Bland-Altman and Correlation plots of (a) Standard non-AI automatic motion correction (MC) vs. average (Avg) manual MC and (b) Deep Learning (DL)-based MC vs. average manual MC for myocardial flow reserve (MFR). The top row depicts Bland–Altman plot and the bottom row shows the correlation plots. CCC: Concordance correlation coefficient, CI: Confidence interval

#### Myocardial Blood Flow

Figure 5a shows BA and correlation plots for standard non-AI based automatic MC and operator average MC. Figure 5b shows the BA and correlation plots between the DL-MC and average MC by two experienced operators for MBF. The variability around the mean difference for DL-MC, ±0.24 (mean difference = 0.00), was lower than variability for non-AI automatic MC, ±0.39 (mean difference = -0.02). Morgan-Pitman resulted in statistically significant difference in variances, with a p-value <0.00001. The bottom row shows correlations between minimal segmental MBF with average of manual-MC from two operators and DL-MC with CCC=0.983 (95% CI: 0.966, 1.000) and non-AI automatic MC with CCC=0.957 (95% CI: 0.930, 0.983). Fisher’s z-score evaluation resulted in statistically significant difference in CCC, p-value <0.00001.

**FIG. 5.**
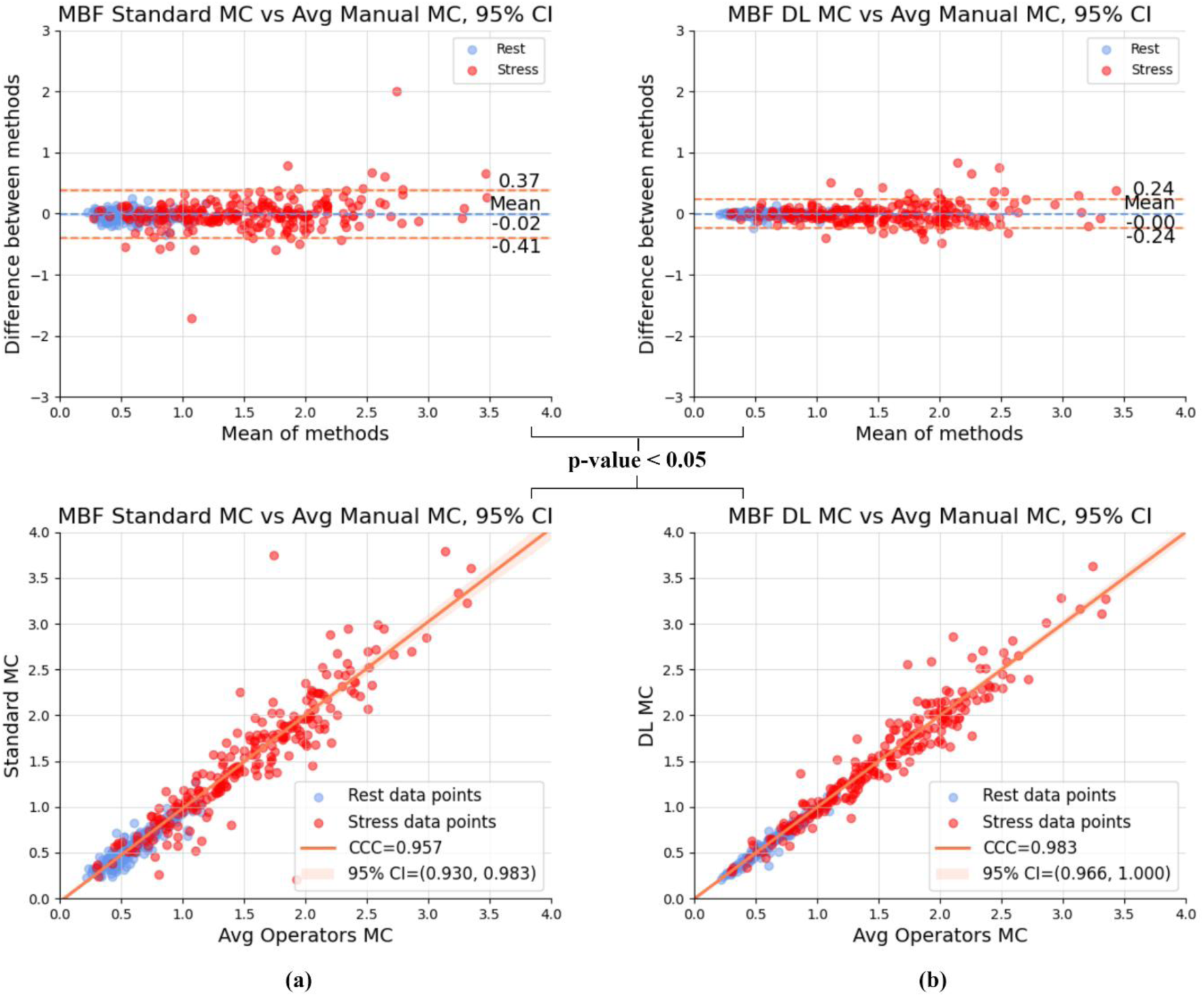
Bland-Altman and Correlation plots of (a) Standard non-AI automatic motion correction (MC) vs. average (Avg) manual MC and (b) Deep Learning (DL)-based MC vs. average manual MC for stress and rest minimal segmental myocardial blood flow (MBF). The top row depicts Bland–Altman plot and the bottom row shows the correlation plots. CCC: Concordance correlation coefficient, CI: Confidence interval

#### Segmental agreement with operators and between operators

To assess segmental performance correlation, CCC was calculated between segmental MFR values obtained from our DL-MC and the average of manual-MC from two operators, as illustrated in Figure 6a. Additionally, CCC was computed between the segmental MFR for manual-MC results from the two operators to evaluate the variability among human operators as shown in Figure 6b. Similarly, Figure 6c and 6d show the CCC for segmental MBF.

**FIG. 6.**
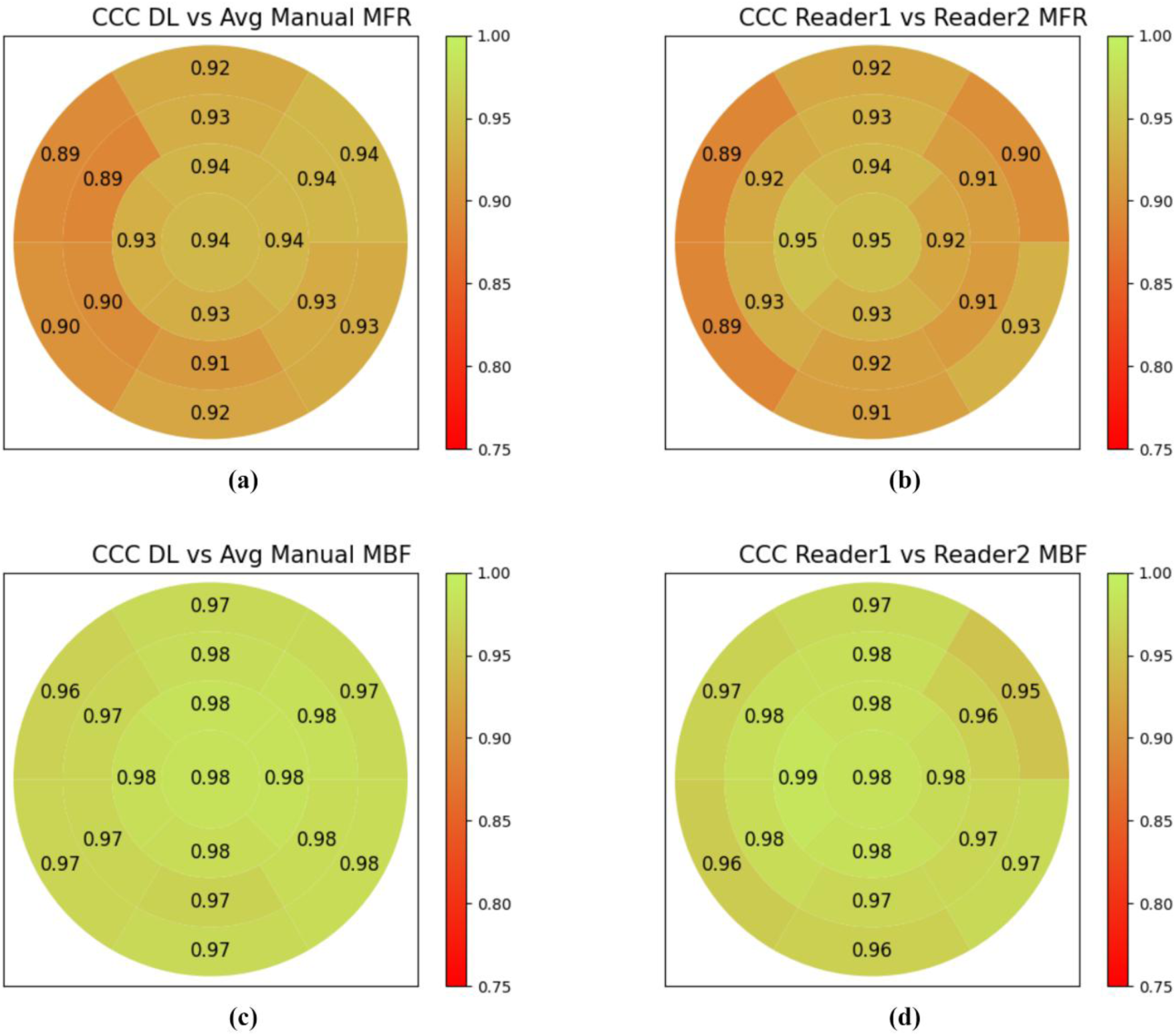
Concordance Correlation Coefficients (CCC) of Deep Learning (DL)-based motion correction (MC) and average (Avg) manual MC for (a) myocardial flow reserve (MFR) and (c) myocardial blood flow (MBF), per segment. Per segment CCC for (b) MFR and (d) MBF with manual motion corrections from two different operators

### Diagnostic Performance

Receiver Operating characteristic (ROC) analysis demonstrates the AUC of minimal segmental stress MBF (Figure 7a). The AUC of MBF with DL-MC saw an improvement over non-AI automatic MC and was significantly greater than no-MC (p<0.05). A strong agreement among the minimal segmental MBF values derived from DL-MC and manual-MC for two operators was noticed for predicting obstructive CAD, with no significant difference (p=0.379 for Operator 1 and p=0.669 for Operator 2).

**FIG. 7.**
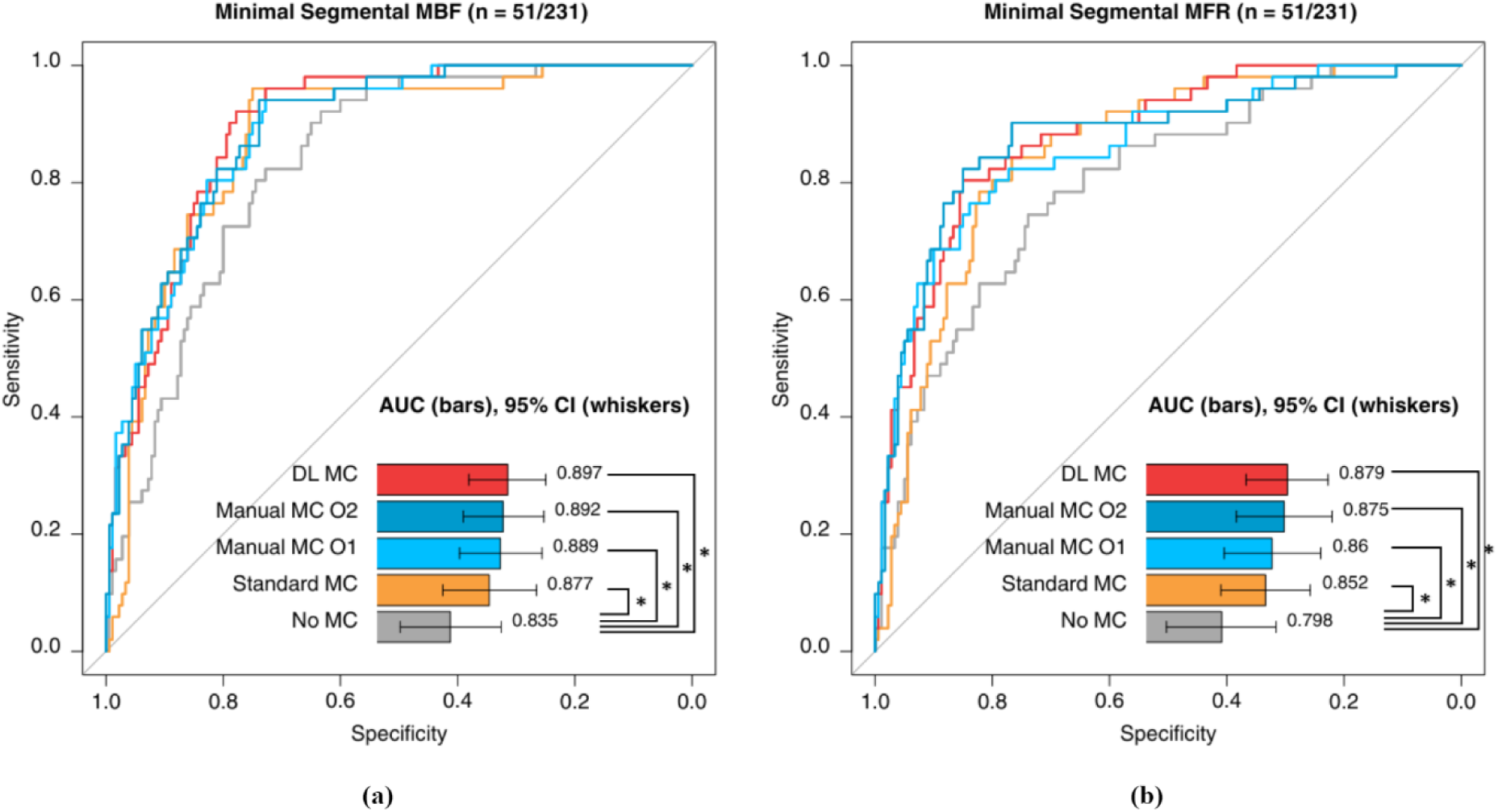
Per-patient diagnostic performance of minimal segmental (a) stress myocardial blood flow (MBF), (b) myocardial flow reserve (MFR) with residual activity correction applied, compared among manual motion correction (MC) from Operator 1 (O1) (light blue), Operator 2 (O2) (dark blue), standard non-AI automatic MC (orange), deep learning (DL) based automatic MC (red) and no-MC (grey). AUC: area under the receiver operating characteristic curve; CI: confidence interval; *: significant (p < 0.05)

Figure 7b depicts the ROC analysis for minimal segmental MFR. The AUC for minimal segmental MFR with DL-MC saw an improvement over non-AI automatic MC and was significantly higher than no-MC (p<0.05). Additionally, there was strong agreement among the minimal segmental MFR values derived from DL-MC and manual-MC readings from two operators for predicting obstructive CAD, with no significant difference observed (p=0.073 for Operator 1 and p=0.754 for Operator 2).

Per-vessel analysis showed that DL-MC was in strong agreement with the manual operators, significantly improved performance in comparison to standard non-AI MC and demonstrated a statistically significant difference in performance compared to no MC (Supplemental Figure 1).

## Discussion

In this work, we conducted an evaluation of the diagnostic performance of stress MBF and MFR calculated using DL-based automatic MC in ^18^F-flurpiridaz PET-MPI, comparing its accuracy to manual operator MC, non-AI automatic MC and no-MC. Manual operator-based motion correction is very time consuming and has inter-operator variability. Hence, we proposed a 3D ResNet based architecture to predict the motion translation vectors. While traditional medical image registration involves comparing highly similar images, in motion correction we explore dynamic image registration which must contend with substantial image variations caused by tracer movement. Therefore, our approach involves generating a multitude of simulated motion vectors for each individual frame using bootstrap distributions of manual motion vectors associated with that frame. Different image pre-processing steps were used to achieve robust motion estimation.

In a previous study, our group demonstrated that RAC and manual-MC improved MBF estimation and CAD diagnosis in ^18^F-flurpiridaz PET-MPI (*7*). A comparison of manual-MC by two expert operators highlights discrepancies between observers, mirroring clinical practice, which bolsters the claim that manual-MC is susceptible to inter-operator variability. Furthermore, the time-intensive manual-MC leads to omission of small shifts by the operators. In contrast, the DL-based automatic MC algorithm offers a standardized approach that reduces subjectivity and observer variability. Despite this, DL-MC achieved comparable or slightly better diagnostic performance for predicting obstructive CAD to manual-MC, indicating its clinical viability. Previous studies using Rubidium-82 PET-MPI have also shown that automatic motion correction enhances test-retest reliability and reduces inter-observer variability, thereby improving reproducibility (*25–29*). This is particularly crucial for the novel Flurpiridaz tracer, as it will be adopted by numerous centers with limited experience in dynamic Flurpiridaz PET imaging.

In another study, our group implemented a standard non-AI automatic MC which improved CAD diagnosis and corrected motion in approximately 10 seconds per case (*8*). Our DL-MC improved the diagnostic performance of minimal segmental stress MBF and MFR for predicting obstructive CAD compared to no-MC and non-AI automatic MC. In comparison to non-AI automatic MC, DL-MC obtained substantially better correlation with average of manual-MC by two operators. Bland-Altman plots demonstrate that DL-MC exhibits significantly lower bias and narrower limits of agreement than non-AI automatic MC, with fewer outliers. The results of this study strongly support the implementation of DL-MC as an essential step in obtaining reliable MBF and MFR measurements for the accurate prediction of coronary artery disease in PET-MPI.

In contrast to other DL-based motion correction algorithms, where only generic simulated motion is used for training and evaluation of the model (*11*), our approach is fundamentally grounded in clinical reality. We use corrections from experienced operators as the ground truth and, crucially, employ a specialized data augmentation strategy. By generating simulated motion vectors from a bootstrap distribution of these real manual corrections, we ensure our model trains on a vast yet realistic spectrum of movements. This is a critical distinction as generic simulations may not accurately mirror the complex and subtle patterns of actual patient motion, whereas our method directly enhances the model’s ability to generalize and perform robustly on real-world clinical data. We also perform site-wise external validation to ensure the model is not biased to a specific mode of acquisition. To account for the limited number of cases with manual correction vectors, each frame of the dynamic PET scan is considered as a sample for our dataset. Additionally, we provide a definitive evaluation including the diagnostic evaluation and comprehensive comparison to experienced operators and standard clinical methods.

Once trained, the network processed individual dynamic image sequences in just 1-1.5 seconds, a 400-fold speedup compared to manual-MC, and over 7-fold speed up compared to non-AI version. As a post-processing tool operating on reconstructed images rather than raw sinogram data, the proposed method can be seamlessly integrated into standard clinical workflows.

### Study Limitations

Our study has several limitations. First, the performance of the network is inherently limited by the quality of the ground truth data used during training. As the manual-MC is susceptible to inter-operator variability, though we used randomized operator targets, errors in targets can propagate through the training process. To mitigate the impact of this issue, we plan to expand our dataset by acquiring additional operator corrections. Second, our method currently focuses on correcting inter-frame motion alone. However, intra-frame motion can lead to distorted or blur frames which may also limit the accuracy of inter-frame MC. Future work will explore and integrate intra-frame motion correction techniques into our network to further enhance its performance. Third, the study population was small (n=231) with a limited number of positive invasive angiography results (n=51). Analyzing the diagnostic accuracy of MC for CAD across different patient subgroups would require a larger sample size and is beyond the scope of this study. Such an analysis could be a valuable focus for future research.

## Conclusion

The 3D ResNet-based architecture trained with manual motion corrections and simulated motion vectors derived from bootstrap distribution of real manual motion showed significant improvement in agreement with experienced operators as compared to standard non-AI automatic MC. The AI-algorithm is significantly faster but diagnostically equivalent to manual-MC and standard automatic MC when corrected MBF and MFR values are used for the detection coronary artery disease.

## KEY POINTS

### QUESTION

Can a deep learning (DL) based motion correction (MC) framework for ^18^F-flurpiridaz PET-MPI improve the efficiency and agreement with expert clinical observers compared to standard non-AI automatic motion correction?

### PERTINENT FINDINGS

In this study, a 3D ResNet-based DL model reduced motion correction time to just 1-2 seconds per patient, while demonstrating excellent agreement with experienced human operators. DL-MC showed equivalent diagnostic performance in predicting obstructive CAD on external evaluation data in comparison to expert clinical observers, with statistically significant improvements in correlation, bias, and limits of agreement with manual-MC compared to standard non-AI automatic method.

### IMPLICATIONS FOR PATIENT CARE

DL-based motion correction could streamline ^18^F-flurpiridaz PET-MPI workflows, improving clinical efficiency while maintaining diagnostic accuracy, ultimately enhancing patient outcomes through faster and reliable detection of obstructive CAD.

## Data Availability

To the extent allowed by data sharing agreements and IRB protocols, the deidentified data and data analysis code from this manuscript will be shared upon written request.

## Disclosures

Dr. Piotr Slomka and Mr. Paul Kavanagh participate in software royalties for QPS software at Cedars-Sinai Medical Center. Dr. Slomka declares an equity interest in APQ Health and has received research grant support from Siemens Medical Systems and consulting fees from Synektik SA and Novo Nordisk. Dr. Buckley is an employee of GE Healthcare. The remaining authors have no relevant disclosures.

### Acknowledgement

This research was supported in part by grant R35HL161195 from the National Heart, Lung, and Blood Institute/ National Institutes of Health (NHLBI/NIH) and R01EB034586 from the National Institute of Biomedical Imaging and Bioengineering (PI: Piotr Slomka). The content is solely the responsibility of the authors and does not necessarily represent the official views of the National Institutes of Health.

