## Supplemental Material for "Dynamic frame-by-frame motion correction for ^18^F-flurpiridaz PET-MPI using convolution neural network"

Study Population

The trial enrolled participants who could undergo either exercise or pharmacologic stress tests, who had undergone clinically indicated myocardial perfusion SPECT and invasive coronary angiography without any prior intervention and were of age 18 years and older(*1*). Exclusion criteria included individuals with a history of percutaneous coronary intervention, myocardial infarction, or other invasive coronary procedures within 6 months of $^{18}$F-flurpiridaz PET. Patients with current non-ischemic cardiomyopathy or a history of coronary artery bypass graft surgery were also excluded (*1*). Each study site obtained approval from their respective Institutional Review Board or ethics committee, and the study protocol adhered to the principles of the Declaration of Helsinki. Informed consent was obtained from all participants in writing prior to the commencement of any study procedures.

PET protocol

PET imaging was performed using PET scanners in 2D or 3D mode after participants fasted for a minimum of 3 hours. The imaging protocol involved a rest/stress acquisition sequence with dynamic PET acquisition initiated after a saline flush of 5-10 mL and an intravenous bolus injection of $^{18}$F-flurpiridaz (2.7 ± 0.2 mCi). Pharmacologic stress was induced using regadenoson, adenosine, or dipyridamole, and was followed by a second dynamic PET acquisition and reconstruction at peak stress after a bolus injection of $^{18}$F-flurpiridaz (5.9 ± 0.3 mCi). The average interval between rest and stress doses was 53 ±11 minutes. PET images were captured over a 15-minute period and underwent rigorous quality control, with reconstruction of PET images with standard corrections applied in predefined time frames.

Standard clinical automatic motion correction

The automatic motion correction (MC) algorithm corrected motion by aligning individual image frames to a static 3D geometrical model of the ventricles, defined by the left ventricle (LV) and right ventricle (RV) contours segmented from the sum of dynamic frames beyond the first 120 seconds using 3D rigid-body translations. Key frames were identified from the time-activity curves where well-defined correspondences between tracer distribution and regions in the geometric model were expected: an LV blood-pool peak frame, an LV blood-pool and myocardium crossover frame, and the end of acquisition frame where counts were concentrated in the LV myocardium only. The key frames were corrected by aligning them to the model through maximization of simple count-based similarity metrics developed for each. The in-between key frames were then corrected by registering them to synthetic reference frames generated from linear blending of neighboring key frames with variable ratios, which were derived from the time-activity curve through maximization of mutual information.

Deep learning-based automatic motion correction

Our network employed a 3D ResNet-based architecture, incorporating a regression head at the end to predict translation vectors. Each Conv3D block contained parametric rectified linear unit (PReLU) activation and batch normalization. Dropout (0.5) was also added to the fully connected layers to reduce overfitting. To train our network, different combinations of loss functions were explored. As the goal was to align each frame to the reference frame, mean square error (MSE) loss between predicted and target motion translation vectors was used ($L_{vec}$). Manual operator motion corrections were performed on interpolated volumes which allow sub voxel translation. To accommodate the same in our training, the volumes were translated using trilinear interpolation. MSE loss was applied between volumes translated using predicted translation and target translation vectors ($L_{vol}$). A combination of weighted ($\lambda$=0.5) $L_{vol}$ and $L_{vec}$ were used to update the network parameters.

Training dataset for AI motion correction

If an operator did not make any corrections, in which case the translation vector from that operator was (0,0,0), the translation vector from the other operator was used as the target. This ensures zero motion translations are chosen as targets only if both operators did not make any correction.

Since our data contained 21-26 dynamic frames and each patient had stress and rest scans, the total number of volumes were around ~9700-12,000. Data augmentation, which involved creating simulated motion vectors, was employed to drastically enlarge and diversify the dataset. Train and test splits were made on patients and not individual volumes ensuring all the dynamic frames belonging to one patient were in the same data split.

Data Preprocessing

PET scans acquired using different reconstruction parameters or from various scanners often exhibit varying appearances and voxel spacing. Consequently, it is essential to standardize and normalize the data before training or inference to ensure consistency and reliability in training and analysis. All volumes were resampled to same voxel spacing and cropped to a fixed size to achieve standardization. The cropped volumes were designed to include the region of interest by obtaining the bounding box around the LV mask while relatively incorporating the other three chambers and minimizing peripheral inclusion. The cropped volumes were normalized using the z-score normalization ($z = x - \mu/\sigma$) method for scaling the intensity. Gaussian blur was applied to the volumes to calculate the mean($\mu$) and standard deviation($\sigma$) used for z-score normalization.

Training and Testing Regimen

Each fold in the five-fold cross-validation process underwent specific data splitting procedures as follows: [1] a training split consisting of 147 or 148 patients used to train the model; [2] a validation split with 37 patients designated for tuning hyperparameters and ensuring no overfitting; and [3] a test split containing 47 or 46 patient scans used to evaluate the method. Results were derived by aggregating outcomes from all five test sets. Each test set included cases from sites that were not included in the network's training phase.

Adam optimizer (*2*) was employed with an initial learning rate of $10^{-3}$ and a weight decay of $10^{-4}$ to optimize network parameters. To prevent overfitting, 'reduce on plateau' scheduler was implemented, reducing the learning rate by a specific factor ($10^{-1}$ in our case) if the validation loss failed to improve for 15 epochs (patience=15). Training continued until the learning rate reached $10^{-6}$ which was the minimum threshold in our experiment, and the model with the lowest validation loss was saved as the best model. Our architecture was implemented using Pytorch deep-learning framework. We trained our model on NVIDIA RTX 4090 GPU. The training approximately took 9 hours, batch size of 16 for a max of 400 epochs with early stopping and 5-fold cross validation.

<u>Myocardial blood flow and Myocardial flow reserve quantification</u>

The estimation of myocardial blood flow (MBF) in each coronary region relied on analyzing tracer uptake kinetics within the initial 90 seconds post-injection, assuming a first-pass extraction fraction of 0.94 (*3*). Rate-pressure product (RPP) was calculated as the product of heart rate and systolic blood pressure. Resting MBF values were then adjusted for RPP using the formula $MBF_{adj} = MBF_{rest} / RPP_{rest} \times 8500$, where 8500 represents the $RPP_{ref}$ value recommended by the Society of Nuclear Medicine and Molecular Imaging Cardiovascular Council and the American Society of Nuclear Cardiology (*4*). Stress and rest MBF values in mL/min/g were computed using parametric polar maps. Based on standard myocardial segmentation (*5*), seventeen segmental stress and rest

MBF values were obtained from the polar map sample (*6*). Stress MBF at each segment was divided by the global rest MBF adjusted for RPP to calculate segmental myocardial flow reserve (MFR). Diagnostic evaluation on 231 patients indicated that minimal segmental stress MBF and MFR had superior diagnostic performance compared to global stress MBF and MFR (*7*). Therefore, minimal segmental stress MBF and MFR were used for diagnostic assessment. All MBF and MFR values were automatically derived in batch mode.

### SUPPLEMENTAL FIGURES

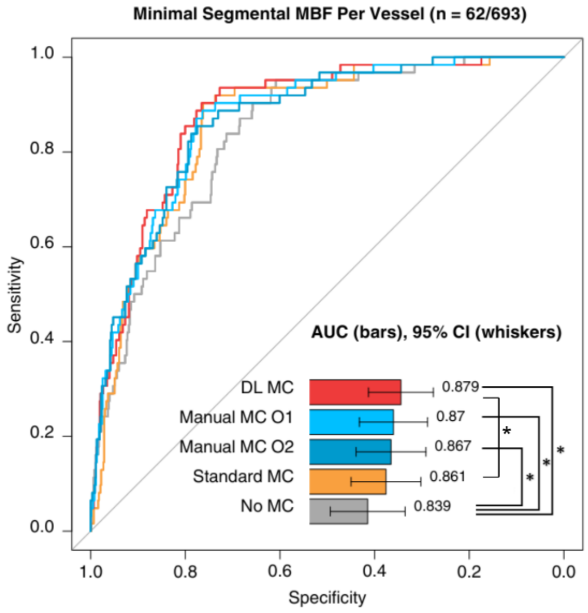


(a)

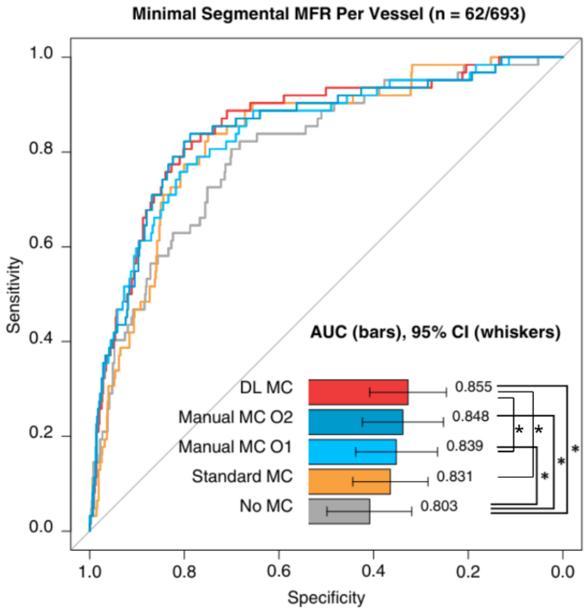


(b)

**SUPPLEMENTAL FIG. 1** Per-vessel diagnostic performance of minimal segmental (a) stress myocardial blood flow (MBF), (b) myocardial flow reserve (MFR) with residual activity correction applied, compared among manual motion correction (MC) from Operator 1 (O1) (light blue), Operator 2 (O2) (dark blue), standard non-AI automatic MC (orange), deep learning (DL) based automatic MC (red) and no-MC (grey). AUC: area under the receiver operating characteristic curve; CI: confidence interval; *: significant ($p < 0.05$)
